# Whole-exome sequencing in children with dyslexia identifies rare variants in *CLDN3* and ion channel genes

**DOI:** 10.1101/2024.12.19.24319320

**Authors:** Krzysztof Marianski, Joel B. Talcott, John Stein, Anthony P. Monaco, Simon E. Fisher, Dorothy V.M. Bishop, Dianne F. Newbury, Silvia Paracchini

**Affiliations:** School of Medicine, University of St Andrews, St Andrews, UK; Institute of Health and Neurodevelopment, College of Health and Life Sciences, Aston University, Birmingham, UK; Department of Physiology, University of Oxford, Oxford, UK; Office of the President emeritus, Tufts University, Medford, MA, USA; Language and Genetics Department, Max Planck Institute for Psycholinguistics, Nijmegen, Netherlands; Donders Institute for Brain, Cognition and Behaviour, Radboud University, Nijmegen, Netherlands; Department of Experimental Psychology, University of Oxford, Oxford, UK; Department of Medical and Biological Sciences, Oxford Brookes University, UK; Centre for Human Genetics, University of Oxford, Oxford, UK

**Author notes:** Correspondence to Silvia Paracchini.

## Abstract

Dyslexia is a specific difficulty in learning to read that affects 5-10% of school-aged children and is strongly influenced by genetic factors. While previous studies have identified common genetic variants associated with dyslexia, the role of rare variants has only recently begun to emerge from pedigree studies and has yet to be systematically tested in larger cohorts. Here, we present a whole-exome sequencing (WES) study of 53 individuals with dyslexia, followed by replication analysis in 38 cases with reading difficulties and 82 controls assessed with reading measures. Our stringent bioinformatics filtering strategy highlighted five brain-expressed genes carrying rare variants: *CACNA1D*, *CACNA1G*, *CLDN3*, *CNGB1,* and *CP*. Notably, a specific variant (7-73769649-G-A) in the *CLDN3* gene was identified in six independent cases, showing a four-fold higher frequency compared to population reference datasets. *CACNA1D* and *CACNA1G* encode subunits of voltage-gated calcium channels (VGCC) expressed in neurons, and variants in both genes have been implicated in neurodevelopmental disorders such as autism spectrum disorder (ASD) and epilepsy. Segregation analysis in available family members were consistent with patterns of dominant inheritance with variable expressivity. In total, high-impact variants in the five genes of interest were found in 26% (N = 14) of individuals of the discovery cohort. Overall, our findings support the involvement of rare variants in developmental dyslexia and indicate that larger WES studies may uncover additional associated genes.

## Introduction

Dyslexia is a neurodevelopmental condition characterised by a specific difficulty in learning to read, occurring in the absence of other causes such as sensory or neurological problems or lack of educational opportunity (1). Regardless of culture and spoken-language, dyslexia affects 5%–10% of children, with a male: female ratio ranging from about 3:1 to 5:1 (2), and often co-occurs with other neurodevelopmental conditions such as developmental language disorders (DLD) and attention deficit hyperactivity disorder (ADHD) (3,4). A strong genetic component for dyslexia is supported by twin studies, which have shown heritability estimates of up to 70%. Moreover, having a first degree relative with dyslexia is the most consistently identified risk factor (5). These observations have motivated molecular studies, but the identification of specific genes has, until recently, been hindered by difficulties in assembling sufficiently powered sample sizes. Lack of national screening programs and heterogeneous assessment and diagnostic criteria pose challenges in recruiting study participants and combining them across studies (1). Initial molecular studies highlighted a handful of genes, including *ROBO1, DCDC2, KIAA0319* and *DYX1C1* (6). However, associations to these genes did not consistently replicate (1).

A breakthrough was made possible by a large (51,800 cases and 1,087,070 controls) genome-wide association study (GWAS) conducted with self-reported dyslexia diagnoses by customers of the direct-to-consumer company 23andMe (7). This study identified 42 independently statistically significant associated loci. About half of these had previously been associated with cognitive phenotypes, while the other half suggested effects more specific to dyslexia. A gene-based analysis reported association with 173 genes. This study also facilitated the generation of dyslexia polygenic scores, which showed significant associations with the reading-related measures available for some cohorts of the GenLang consortium (https://www.genlang.org/), an international project aimed at dissecting the genetics of language-related traits, including reading abilities. An independent quantitative meta-GWAS of reading- and language-related measures in the GenLang cohorts (N ∼34,000) identified only a single genome-wide significant locus but highlighted significant genetic correlations across language-related cognitive domains and traits derived from neuroimaging data (8). The SNP-based heritability from these GWAS efforts ranged between 0.13 and 0.26. Although larger GWAS are expected to identify additional common variants associated with dyslexia, it is likely that rare variants, *e.g.* those with a minor allele frequency (MAF) < 1%, will also play a role.

Whole-exome (WES) and whole-genome sequencing (WGS) studies have demonstrated the role of rare variants in most complex traits (9) including for neurodevelopmental conditions like autism spectrum disorder (ASD) (10), schizophrenia (11), bipolar disorder (12) and childhood apraxia of speech (CAS) (13–15). Rare variants have been reported also for conditions that frequently co-occur with dyslexia, such as ADHD (16) and language impairment (17–19). Rare variant analyses and WES screenings focussing specifically on dyslexia are limited. *DYX1C1*, the first risk gene reported as a dyslexia candidate was found to be disrupted by a translocation in one individual (20). Segregation analyses in large pedigrees suggested the role of variants in the *CEP63* (21)*, NCAN* (22), and *SEMA3C* (23) genes. Large population-based cohorts like the UK Biobank, for which WES data have been generated at scale, are not well-suite for genetic studies of dyslexia because data on dyslexia diagnosis are not reliable and reading-related measures have not been collected (1). Furthermore, it has been shown that rare variants detected in clinical cohorts might not show signals in population samples because of incomplete penetrance *(i.e.* when not all individuals with the variant exhibit the trait) or variable expressivity (when different degrees of a phenotype are associated with the variant) (24).

In the current proof-of-principle WES study, we analysed 53 unrelated individuals with dyslexia. Replication analysis was performed in an independent cohort of 38 cases with reading difficulties (RD) and 82 typically developing (TD) controls, assessed with the same quantitative test as part of a twin study on language-related problems and co-occurring conditions. Our findings highlight rare variants in five genes *CACNA1G, CACNA1D, CLDN3, CNGB1,* and *CP*. Notably, a specific *CLDN3* variant was identified in six independent cases and was not observed in the controls. The availability of family members and quantitative assessments facilitated segregation analysis and enabled the evaluation of the impact of variants on reading-related measures.

## Material and Methods

### Discovery cohort

We analysed a discovery sample of 53 unrelated cases selected from two existing cohorts recruited for genetic studies of dyslexia and which have been previously described (Table 1) (25). The first cohort includes 290 families (689 siblings, 580 parents), most with at least one child diagnosed with dyslexia and a second child presenting reading difficulties. The second cohort comprises 592 singletons with dyslexia or reading problems. Both cohorts were recruited in the South of England at Dyslexia Research Centre clinics in Oxford and Reading or the Aston Dyslexia and Development Unit in Birmingham. All children (probands and their siblings) were individually assessed with the following core tasks: single-word reading (SWR) and single-word spelling accuracy (SPELL)(26), single non-word reading (NWR) (27), phonological awareness (PA) (28), irregular word reading (IWR) (27), orthographic coding (ORTH) (29), as well as measures of verbal (VIQ) and performance (PIQ) IQ (30) (See supplementary Table 1). Parents in the family cohort were not formally assessed but some of them provided information about their own and their family’s history of dyslexia or reading problems by questionnaire during an interview. In both cohorts, probands were excluded if they had been formally diagnosed with co-occurring developmental conditions such as language impairment, autism or ADHD.

**Table 1.** Descriptive statistics of the cohorts.

| Cohort | N (M) | Age | PIQ | SWR | NWR | SPELL | IWR | ORTH | PA |
| --- | --- | --- | --- | --- | --- | --- | --- | --- | --- |
| Discovery | 53 (40) | 12 | 0.17 | -2.04 | -1.52 | -2.30 | -2.39 | -1.79 | -1.25 |
| Replication |  |  | PIQ | SWR | NWR | ACC | COMP | RR |  |
| RD | 38 (22) | 8 | -0.10 | -0.71 | -0.80 | -1.05 | -1.00 | -0.67 |  |
| TD | 82 (36) | 8 | 0.47 | 0.97 | 0.73 | 0.34 | 0.27 | 0.62 |  |
| T-stat |  |  | 3.77 | 11.21 | 10.23 | 11.23 | 10.12 | 8.22 |  |
| *P* |  |  | 1.55E-3 | 1.61E-19 | 3.50E-17 | 2.02E-19 | 8.84E-17 | 2.09E-12 |  |

Phenotypic scores are represented as mean z-scores, which have been standardized based on population distributions with mean=100 and SD=15. N: sample size; M: male; Age: mean age at assessment in years; PIQ: performance IQ, SWR: single word reading; NWR: non-word reading; SPELL: single word spelling accuracy; IWR: irregular word reading; ORTH: orthographic coding; PA: phonemic awareness; ACC: NARA-II accuracy; COMP: NARA-II comprehension; RR: NARA-II reading rate; T-stat: absolute t-test statistic; P-value: Bonferroni adjusted for six tests.

For this study, individual cases were prioritised based on low SWR scores and availability of a high-quality DNA sample. In total, 53 unrelated cases were selected, 33 from the family cohort and 20 from the singleton cohort. WES data were also generated for available family members (N = 14) of variant carriers in the top five genes for segregation analysis.

The study was approved by the Oxfordshire Psychiatric Research Ethics Committee and the Aston University Ethics Committee, with informed consent obtained from all the participants and/or their caregivers.

### Replication and control cohorts

The replication and control cohorts were derived from a UK twin cohort (N = 194 twin pairs) recruited to study language difficulties (31). From this cohort, replication cases and controls were selected based on the following criteria.

Replication cases with reading difficulties (RD) were required to have a PIQ above or equal to −1 SD and at least one score more than 1 SD below the expected level on any of the following tests: single word reading (SWR), non-word reading (NWR) (32), text reading accuracy (ACC), comprehension (COMP) or reading rate (RR) from the NARA-II (33). See Supplementary Table 1 for more details on the tests.

Typically developing (TD) controls were selected based on a PIQ of no more than 1 SD below the expected level for their age and a SWR score above the mean. Children with a Development and Well-Being Assessment (DAWBA) diagnosis of ASD (34) were removed from both groups.

If both twins in a pair met the criteria for inclusion, one twin was randomly selected for the study. This process resulted in a replication group of 38 cases with RD and 82 TD controls with good reading scores. Descriptive statistics and mean phenotypic scores for these two groups are shown in Table 1. WES and phenotypic data were available for their twin siblings for follow up analyses. The study was approved by the Berkshire NHS Research Ethics Committee.

Siblings and family members of the dyslexia and RD cases were assigned to a category of mild reading difficulties if they scored below 1 SD on at least one reading measure or had an average score across all reading measures at least 0.25 SD below the mean.

### Bioinformatics analysis

WES data for all samples were generated using Illumina technology (NovaSeq 6000, Q30 80%, with 50X coverage). Raw sequences were trimmed using Trimmomatic (35). Trimmed reads were mapped to the human reference genome (GRCh38) using bwa-mem (https://github.com/lh3/bwa). Picard tools (http://broadinstitute.github.io/picard/) were used for read-groups replacement, removal of PCR duplicates, and base recalibration. The pre-processed reads were indexed using SAMtools (https://www.htslib.org/) and called with DeepVariant v1.4.0 (36). VCF files were annotated with ANNOVAR (37) to identify high-impact variants.

In accordance with the ACMG guidelines (38), high-impact variants were defined as rare variants (PM2) predicted to damage protein function (PP3). Accordingly, variants were removed if they had a minor allele frequency (MAF) ≥ 1% (based on gnomAD v3.1.2 for Non-Finnish Europeans), or if they were annotated as synonymous, intronic or intergenic variants. To be retained, variants had to be predicted as damaging by all five predictive algorithms (SIFT, PolyPhen2, LRT, MutationTaster and FATHMM) or already annotated as pathogenic or likely pathogenic in ClinVar for any disorders (https://www.ncbi.nlm.nih.gov/clinvar/).

Genes were selected as candidates if high-impact variants were detected in three or more independent cases in the discovery cohort (in accordance with Genomics England guidelines (https://panelapp.genomicsengland.co.uk/). To warrant further follow-up, the gene also had to carry at least one high-impact variant in the RD replication cases and no such variants in the TD replication controls.

Genomic localisation of the variants of interest was based on UniProt and InterPro data. Exonic locations were accessed from UCSC Table Browser hg38.refGene.

### Post-WES analyses

The overlap between the genes identified as candidates in the discovery cohort and those found to be associated in the dyslexia GWAS by Doust et al. (2022) was tested with the GeneOverlap package in R (https://rdrr.io/bioc/GeneOverlap/).

Gene-set enrichment was performed in FUMA (GENE2FUNC tool https://fuma.ctglab.nl/) using Gene Ontology classifications and expression data from GTExv8.0. A list of unique genes was compared against all protein-coding genes, with Benjamini-Hochberg (false discovery rate, FDR) correction applied for multiple testing. Enrichment within brain regions at different developmental stages was tested using the BrainSpan dataset.

Brain expression levels for top genes were extracted from GTEXv8.0 (39) and reported as the maximum observed normalised gene expression across 13 brain regions: amygdala, anterior cingulate cortex, basal ganglia, cerebellar hemisphere, cerebellum, cortex, frontal cortex, hippocampus, hypothalamus, nucleus accumbens, putamen, spinal cord, and substantia nigra.

## Results

WES analysis in this discovery cohort (N=53) identified 580 high-impact variants (as defined in Methods) across 512 genes (Supplementary Table 2). On average, each individual carried 11 high-impact variants.

To investigate overlaps with previous findings from analyses of common variants, we compared the identified genes with the 173 genes reported in gene-based analysis of the dyslexia GWAS by Doust and colleagues (7). There was no statistically significant overlap (P = 0.16) and only seven of the GWAS-identified genes carried high-impact variants in our current study: *CUX2, HSPB2, INA, MITF, SCN5A, SGCD,* and *WDR38*. Among these, *INA* was the only gene that carried high-impact variants in more than one independent case (N = 2; Supplementary Table 2).

The 512 genes identified in our analysis did not include any previously reported candidate genes associated with dyslexia through common (*i.e. KIAA0319* and *DCDC2)* or rare (*i.e. CEP63, DYX1C1, NCAN, ROBO1,* and *SEMA3C*) variants. However, our annotations for disease associations highlighted two different variants in *ZGRF1,* a gene previously implicated in CAS (Peter et al., 2016), in two independent cases. Gene pathway enrichment analysis showed that the top associated pathways were action potential for the biological processes (BP, *p* _adj_ = 3.55E-6) category, membrane protein complex for the cellular component (CC, *p* _adj_ = 5.23E-14) category, and adenyl nucleotide binding for the molecular function (MF, *p* _adj_ = 4.56E-14) category (Supplementary Tables 3–5).

Analyses of gene expression profiling data did not show enrichment for brain expression in any specific regions when testing GTEx v8 general tissue types (Supplementary Table 6) or BrainSpan datasets (Supplementary Tables 7 and 8).

There were 22 genes that presented high-impact variants in at least three independent cases (Supplementary Table 9). The gene with the highest number of high-impact variants was *CFTR*, with six different variants identified in seven individuals. This was followed by *CLDN3* which had the same 7-73769649-G-A variant reported in five unrelated individuals.

The top associations in gene pathways enrichment analysis for these genes were membrane depolarization (BP, *p* _adj_ = 0.01; Supplementary Table 10), transporter complex (CC, *p* _adj_ = 4.26E-3; Supplementary Table 11), and high voltage-gated calcium channel activity (MF, *p* _adj_ = 2.63E-3; Supplementary Table 12). This group of genes did not exhibit specific patterns of brain expression (Supplementary Tables 13–15).

We then conducted a replication analysis for the 22 genes in 38 individuals with reading difficulties (RD replication cases) and in 82 typically developing (TD) controls (Table 1). In the RD replication cases, a gene was considered replicated if at least one high-impact variant was present, even if this involved a different variant from that identified in the discovery cohort. Additionally, our replication criteria required that no variants identified in the discovery and RD cases could be present in the TD group. Under these criteria, five genes showed evidence of replication: *CACNA1D*, *CACNA1G*, *CLDN3*, *CNGB1* and *CP*. An additional 7-73769649-G-A rare allele in *CLDN3* was identified in the RD cases (Table 2).

**Table 2.** Genes meeting our prioritisation criteria.

| Gene | Variant | gnomAD AF | CADD score | Discovery<br>N=53 | Replication<br>N=38 | Control<br>N=82 | Max brain<br>Expression** | Nervous system<br>region |
| --- | --- | --- | --- | --- | --- | --- | --- | --- |
| **CLDN3** | 7-73769649-G-A | 0.0078 | 33 | 5 | 1 | 0 | 2.03 | Cerebellar<br>Hemisphere |
|  | **All variants** |  |  | **5** | **1** | **1\*** |  |  |
| **CACNA1G** | 17-50600739-G-A | 0.0034 | 28 | 3 | 1 | 0 | 3.86 | Cerebellum |
|  | 17-50601149-G-A | 0.000031 | 34 | 1 | 0 | 0 |  |  |
|  | **All variants** |  |  | **4** | **1** | **0** |  |  |
| **CACNA1D** | 3-53747393-G-A | 0.0000155 | 34 | 1 | 0 | 0 | 1.7 | Cerebellum |
|  | 3-53749301-G-T |  | 28.8 | 1 | 0 | 0 |  |  |
|  | 3-53751789-G-A | 0.0000465 | 34 | 1 | 0 | 0 |  |  |
|  | **All variants** |  |  | **3** | **1\*** | **0** |  |  |
| **CNGB1** | 16-57897524-C-T | 0.0061 | 34 | 2 | 1 | 0 | 2.48 | Hypothalamus |
|  | 16-57901371-T-A | 0.0011 | 31 | 1 | 0 | 0 |  |  |
|  | **All variants** |  |  | **3** | **2\*** | **0** |  |  |
| **CP** | 3-149178609-C-G | 0.0023 | 27.4 | 1 | 1 | 0 | 0.87 | Spinal cord |
|  | 3-149199783-G-A | 0.0042 | 28.3 | 1 | 0 | 0 |  |  |
|  | 3-149207611-T-C | 0.0006 | 25.8 | 1 | 0 | 0 |  |  |
|  | **All variants** |  |  | **3** | **1** | **1\*** |  |  |

\* variant in the same gene but different from those identified in the discovery cohort; \*\* maximum observed normalised gene expression across 13 brain regions; CADD = Combined Annotation Dependent Depletion.

For three genes (*CACNA1G*, *CACNA1D*, and *CNGB1)*, high-impact variants tended to cluster in adjacent exons and, consequently, in spatially close regions of the resulting protein (Figure 1).

**Figure 1.**
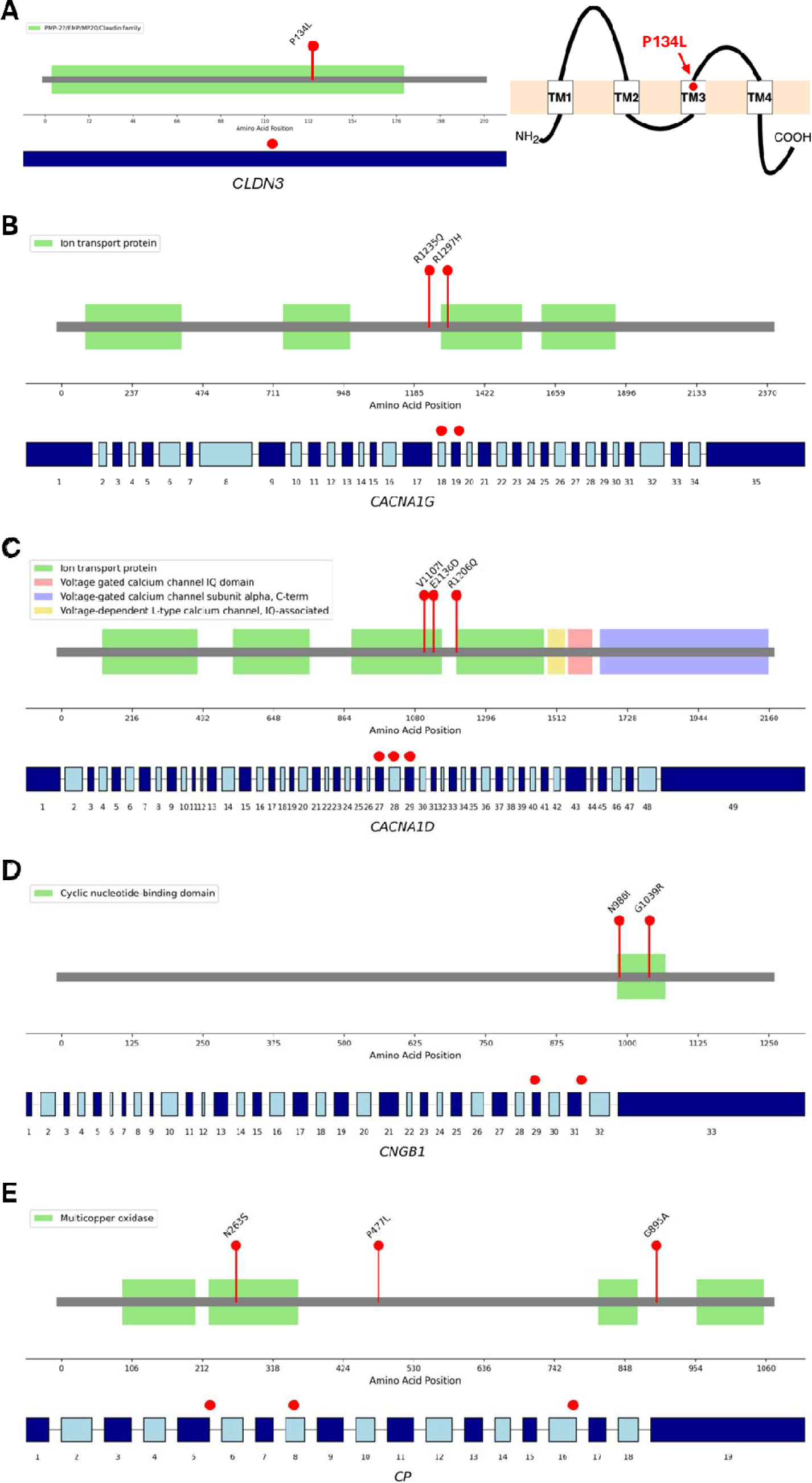
Schematic showing high-impact variants in five replicated genes. The location of high-impact variants (red dots) is shown in relation to exons (lower tracks) and protein domains (upper track) for the A) *CLDN3*, B) *CACNA1G*, C) *CACNA1D*, D) *CNGB1* and E) *CP* genes. A) A 2D structure of the single-exon CLDN3 gene shows that the 7-73769649-G-A variant leads to the P134L amino acid change in the third transmembrane (TM3) domain of the protein.

All high impact variants occurred in heterozygous status. One individual carried high impact variants in three of these genes (*CNGB1, CLDN3* and *CACNA1D)* and two individuals carried high impact variants in two genes (*CACNA1D* and *CLDN3; CACNA1G* and *CLDN3)*.

We compared reading scores between the carriers and non-carriers of the *CLDN3* variant which showed highest recurrence in the discovery cohort (Supplementary Table 16). On average the carriers of this variant exhibited lower scores on all six reading-related measures. The individual in the RD replication cases who carried the 7-73769649-G-A rare allele exhibited a high PIQ (z-score = 1.13), an extremely low reading scores (*i.e.* SWR z-score = −3; average reading z-scores = −2.23). Their dizygotic twin sibling shared the same variant and did not meet criteria for assignment to either the RD or the TD group. This sibling presented a high PIQ score (z-score = 1.6) and all five reading scores were below the mean (average z-score = −0.68), meeting criteria for mild reading problems.

Finally, we evaluated the segregation patterns of variants in the five replicated genes, for cases in the discovery cohort where family members were available. Siblings had been assessed using the same battery of tests as the cases. Parents had not been assessed systematically, but for some of them self-reported information for dyslexia diagnoses or reading difficulties were available.

No data on family members were available for carriers of variants in the *CACNA1G* and *CP* genes. Only one family was available for *CNGB1* (16-57901371-T-A), two families were available for *CLDN3* (7-73769649-G-A), and one family had variants in three genes (*CNGB1* 16-57901371-T-A, *CLDN3* 7-73769649-G-A and *CACNA1D* 3-53751789-G-A; Figure 2).

**Figure 2.**
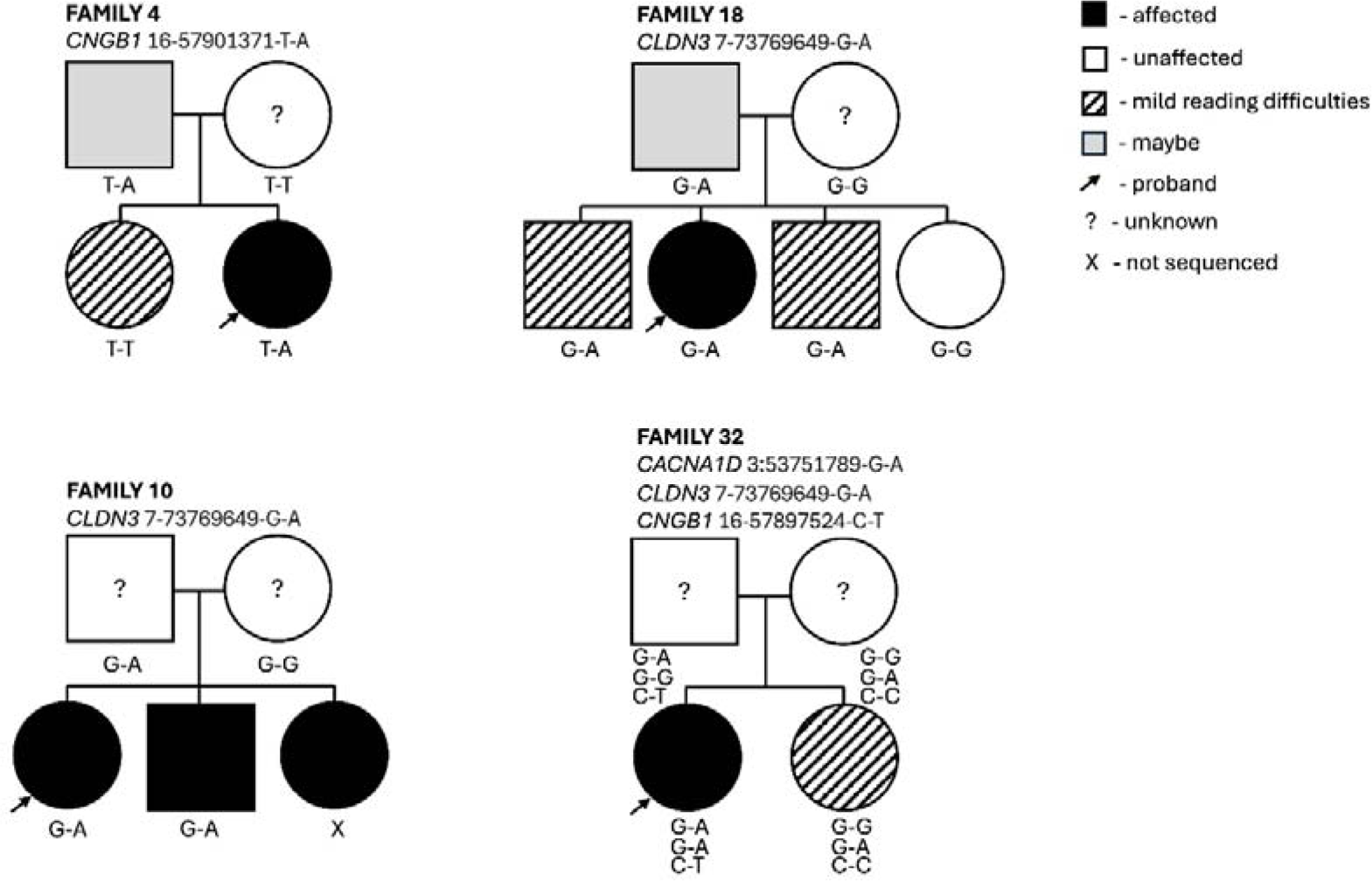
Segregation analysis. Four families were available for segregation analysis of the variants in the five candidate genes. Overall, segregation patterns are consistent with dominant effects with variable expressivity. The phenotypes of the probands and their siblings were defined through a battery of cognitive tests aimed at assessing reading abilities. Parents were not formally assessed, and their phenotypic status was based on self-reported information. Parents who answered “maybe” when asked if they had dyslexia are indicated in grey. A question mark indicates that no information was available for the parents. Mild reading difficulties (striped pattern) in the children were defined as having either at least one of the reading scores lower than 1 SD from the mean or an average score across all reading measures at least 0.25 SD below the mean.

Overall, the observed segregation patterns were consistent with dominant effects with variable expressivity as all individuals carrying high-impact variants and for whom data were available exhibited some form of reading difficulties.

## Discussion

We report the results of a WES study of dyslexia conducted in a discovery cohort (N = 53) and followed up in a replication cohort (N = 38 RD cases and N = 82 TD controls), representing the largest analysis of this kind conducted to date. All cases and their siblings were assessed in person with a battery of standardised tests. Our findings highlight five replicated genes with high-impact variants detected in 26% (N = 14) individuals of the discovery cohort.

The observation with highest recurrence was a specific rare variant (7-73769649-G-A) in the CLDN3 gene which was observed in a total of six unrelated cases across the discovery and RD cohorts (N_total_ = 91). This corresponds to an allele frequency of 3.3%, representing a four-fold increase compared to the frequency of 0.8% reported for this variant in the gnomAD samples of European ancestry. Moreover, this variant was not observed in the TD controls, minimising a potential population structure effect caused by a recruitment bias within the UK.

CLDN3 is a single exon gene spanning 1274 bases in total at 7q11.23, within the critical region deleted in Williams syndrome, a developmental disorder characterized by learning difficulties, distinctive facial features, and cardiovascular problems (41). This gene encodes Claudin-3, a protein expressed in different epithelia with roles for the formation of tight junctions and the blood-brain-barrier (42). Depletion of Cldn3 has been shown to lead to neural tube defects using a chick embryos model system. Targeted analysis of CLDN genes in 152 patients with spinal neural tube defects (NTDs) identified eleven variants, including a A128T missense variant in CLDN3. Overexpression of the CLDN3 A128T variant in chick embryos resulted in a significant increase in NTDs in this model (43). The 7-73769649-G-A variant identified in this study results in a P134L amino acid change, which, like A128T, is located in the third transmembrane (TM3) domain of Claudin-3. In both Claudin-3 and Claudin-5, the TM3 domain is critical for tight junctions assembly, as demonstrated by studies in HEK293 cells (44). These observations, combined with a high CADD score (= 33; Table 2), predict that the 7-73769649-G-A variant is likely to impact the function of the CLDN3 protein.

Among the five replicated genes were two members of the voltage-gated calcium channel (VGCC) family, CACNA1D and CACNA1G. VGCC genes are critical for proper brain function, mediating essential calcium-dependent processes such as gene transcription, neurotransmitter release, and neurite outgrowth (45). Both CACNA1D and CACNA1G, which show high level of expression in the cerebellum, have been previously implicated in neurodevelopmental disorders. CACNA1D variants have been associated with ASD, global developmental delay and other neurodevelopmental conditions (46,47). CACNA1G variants have been associated with infantile-onset syndromic cerebellar ataxia (48), severe neurodevelopmental delay, epilepsy (47) and intractable seizures (49). Furthermore, we observed high-impact variants in another VGCC gene, CACNA1C, in two independent cases from the discovery cohort (Supplementary Table 2). No variants in CACNA1C were observed in the TD group.

For the remaining two replicated genes, there is less evidence for a role in neurodevelopment. The CNGB1 gene encodes the cyclic nucleotide-gated channel beta 1 protein, which is a subunit of the rod photoreceptor cyclic nucleotide-gated channels. These channels ensure a flow of ions into the rod photoreceptor outer segment in response to light-induced changes in intracellular cyclic GMP levels. CNGB1 variants are associated with retinitis pigmentosa, a degenerative eye disease that affects photoreceptor function (50). CP encodes ceruloplasmin, a major copper-carrying protein in the blood. Variants in the CP gene can lead to aceruloplasminemia, a rare genetic disorder characterised by iron accumulation in the brain and other organs which can result in neurological and systemic symptoms (51). Elevated levels of iron in the cortical speech motor network have been reported in people who stutter (52).

In terms of intragenic localisation, the variants detected for CACNA1D, CACNA1G and CNGB1 clustered within restricted regions of each gene. Specifically, the three CACNA1D (one novel and two ultra-rare) variants, cluster within exons 26-28, while all two CNGB1 variants cluster within the cyclic nucleotide-monophosphate (c-NMP) binding domain (Figure 1).

We did not detect variants in genes previously identified in GWAS, WES, or candidate gene association studies related to dyslexia, and none of the five genes highlighted in our study have been previously implicated in dyslexia. This underscores the highly polygenic nature of the condition.

However, our annotations highlighted two unrelated individuals with distinct variants in ZGRF1 (Supplementary Table 2), one of which was the same variant (4-112585555-C-T) previously reported in CAS, a language-related condition (40). The same 4-112585555-C-T variant was present also in a TD control and therefore studies of larger cohorts are required to clarify the role of this gene in language-related conditions.

The gene with highest number of recurrent variants observed in the discovery cohort was CFTR, which was not retained as a gene of interest because one of the variants detected in the discovery sample was also present in the TD group. Variants in CFTR are known to cause cystic fibrosis, and its role in neurodevelopment is not well established. However, early studies reported a significant linkage signal for language impairment at the CFTR locus (53). These two examples highlight the importance of including controls with relevant phenotypic measures to highlight potential false positives. Conversely, the likelihood of false negatives was increased by the relatively small size of our sample, requiring stringent filtering criteria for pathogenicity predictions. WES analyses in larger cohorts will enable a more systematic evaluation of the role of rare variants to dyslexia, such as for the spectrum of CFTR variants.

The detailed cognitive assessment of cases and their family members allowed us to further evaluate the phenotypic effects of the top five genes and variants. Although limited to four families, the segregation patterns were consistent with dominant effects for the three genes that were analysed. All variant carriers with available phenotypic data exhibited some degree of reading problems. In family 4, the CNGB1 risk variant is transmitted from the father, who self-reported reading problems, to the proband but not to the sibling. The sibling presents with mild reading difficulties suggesting that other risk factors contribute might be present in this family. The patterns in family 10 and 18 are consistent with a dominant effect of the CLDN3 7-73769649-G-A variant on dyslexia and/or reading difficulties. In family 18, the variant is transmitted from the father with self-reported reading difficulties to three siblings with either dyslexia (PIQ z-score = 1.5; SWR z-score = −2.2; average reading measures z-score = −2.51) or mild reading problems. One of the siblings with a mild phenotype has a high PIQ (z-score = 1.7) and an average z-score across the reading measures of −0.54. The other sibling does not have a PIQ measure but presents with a high verbal IQ (z-score = 1.6) and an average z-score across the reading measures of −0.4. The fourth sibling does not have the variant and is a good reader (SWR z-score = 1.3). Finally, in family 32, the proband carries three risk variants and presents exceptionally low reading scores (average across six reading tasks = −2.5 SD) despite having a PIQ above the mean (z-score = 0.1). The sibling, who shares only the CLDN3 variant out of the three, presents a mild phenotype characterised by a high PIQ (z-score = 2.3) and low z-scores on specific reading tasks (IWR = −0.61, ORTH = −1.16).

A large discrepancy between PIQ and reading scores were observed for the carrier of the CLDN3 variant identified in the RD replication cases (PIQ z-score = 1.13; average reading z-scores = −2.23) and the carrier’s sibling (PIQ z-score = 1.6; average reading z-scores = −0.68) who shared the same variant. Overall, such patterns support a role for these rare variants in dyslexia and varying degrees of reading difficulties.

The phenotypic variability suggests that additional factors modulate the effects of these variants. It is worth nothing that, in the discovery cohort, the other two carriers of the CLDN3 variant for whom we had no family data, also carried a high impact variant in either CACNA1D or CACNA1G. This suggests that the effect of the CLDN3 variant on reading abilities could potentially be modulated by other rare risk variants. The phenomenon of variable expressivity, as well as incomplete penetrance, has been reported for most complex traits, including psychiatric and cognitive phenotypes (24). These phenomena are likely to reflect the interaction of genetic, environmental, and lifestyle factors in influencing phenotypic outcomes. Typically, rare variants discovered in clinical cohorts with specific phenotypic characteristics tend to be associated with milder manifestations in the general population, likely due to the absence of such interacting risk factors.

In conclusion, this study advances our understanding of potential roles of rare genetic variants in dyslexia. We identified five novel candidate genes. These include CLDN3 which plays several key functions in neurodevelopmental processes. Notably, a specific variant in CLDN3 was identified in six independent cases. Two other genes, CACNA1D and CACNA1G, members of the VGCC gene-family, are involved in neuronal excitability, and have previously been linked to other neurodevelopmental conditions. While our findings do not overlap with previously reported dyslexia-associated genes, they align with the established role of VGCC genes in psychiatric and neurodevelopmental conditions. We extend the significance of this pathway to dyslexia, suggesting that VGCCs may contribute to its aetiology and linking these genes to shared mechanisms across neurodevelopmental traits. Additionally, genes such as CFTR and CACNA1C, which were excluded based on our stringent filtering criteria, may represent potential candidates for future investigations.

These findings underscore the importance of systematic WES or WGS analysis in larger, well-characterised cohorts to identify additional genetic factors and further advance our understanding of the neurobiology underlying dyslexia.

## Code availability

Code used in the current study to analyse the data can be found at https://github.com/kmarianski/DyslexiaWES_CLDN3

## Data Availability

All bioinformatic data produced in the present work are contained in the manuscript or in the supplementary material. The code used to analyse is avaialble online.

https://github.com/kmarianski/DyslexiaWES_CLDN3

## Acknowledgements

We would like to thank the participants, their families, recruiters and health-care staff involved in collection of the data.H

